# No maternal health without mental health: suggested indicators to monitor perinatal mental health globally

**DOI:** 10.1101/2024.10.04.24314919

**Authors:** Francesca Palestra, Allisyn C. Moran, Neerja Chowdhary, Tatiana Taylor Salisbury, Tarun Dua, Shanon McNab, Elly Layton, Elissa C Kennedy, Caroline SE Homer, Jane Fisher, Simone Honickman, Zelee Hill

**Affiliations:** Department of Maternal Newborn Child and Adolescent Health and Ageing (MCA), World Health Organization (WHO), Geneva, Switzerland; Department of Mental Health, Brain Health and Substance Use (MSD), World Health Organization (WHO), Geneva, Switzerland; Centre for Global Mental Health, King’s College London, London, United Kingdom; MOMENTUM Country and Global Leadership, Jhpiego, Washington, District of Columbia, United States of America; Burnet Institute, Australia; Public Health and Preventive Medicine, Monash University, Melbourne Australia; Department of psychiatry and mental health, University of Cape Town, Cape Town, South Africa; Institute for Global Health, University College London, London, United Kingdom

**Keywords:** Perinatal mental health, maternal health, newborn health, depression, mental health conditions, health monitoring, indicators

## Abstract

Perinatal mental health refers to the psychological wellbeing of women during pregnancy and up to one year postpartum. Perinatal mental health conditions as depression significantly affect maternal, newborn health, and child development worldwide. However, the absence of standardized indicators limits effective monitoring and evaluation. This paper introduces a framework with indicators for global perinatal mental health monitoring.

The framework development involved a scoping review, expert consultations, and stakeholder surveys. A global expert group, guided by evidence and the World Health Organization (WHO), identified an initial set of indicators. Two rounds of online surveys were conducted, allowing stakeholders to rank these indicators and suggest additional ones. Indicators were chosen based on their validity, reliability, relevance, feasibility, and potential impact for advocacy purposes on perinatal mental health. The WHO expert working group finalized the set of indicators.

Six perinatal mental health indicators were identified for future testing. These include three primary indicators: policy presence, screening coverage, and prevalence of perinatal mental health conditions. Three secondary indicators cover mental health expenditure, healthcare professional training, and provision of care.

Challenges to implementing standardized monitoring include resource limitations and data collection methods. Strengthening health workers and facility capacity to measure, report, and interpret perinatal mental health data, including the use of screening tools, is crucial. The integration of these indicators into existing systems, such as health information management systems (HIMS) and national surveys, will be key.

Monitoring perinatal mental health is essential for improving maternal and newborn outcomes. The proposed framework offers a means to enhance global monitoring and guide policy interventions. Global stakeholders are encouraged to integrate perinatal mental health into their policy, program, and monitoring agendas.

## Introduction

Perinatal mental health (PMH) is one of the most neglected issues in maternal health [1]. PMH refers to the psychological wellbeing of individuals during pregnancy and up to one year postpartum [2]. This is not to be confused with the perinatal period of a newborn which is seven days after birth [3].

A variety of mental health conditions may be experienced during the perinatal period, including but not limited to depression, anxiety, psychoses and alcohol and substance use [4, 5]. Perinatal mental health conditions pose a significant burden globally, affecting millions of women each year. Globally it is estimated that 10-20 % of pregnant women and those who just have given birth experience depression and rates are highest in low- and middle-income countries [8, 9], Different cultural practices, poverty, biological and social factors have been associated with influencing women’s vulnerability to perinatal mental health conditions [10].

Poor PMH not only has adverse effects on the woman’s quality of life but also impacts the health outcomes of newborns and children [11]. For example, perinatal depression has been linked to suicide and adverse birth outcomes, such as preterm birth and low birth weight, as well as long-term developmental and behavioral issues in children [12, 13]. Perinatal mental health is therefore crucial for the wellbeing of women and their babies.

Despite the prevalence and impact of perinatal mental health conditions, there is a lack of standardized indicators for global monitoring. No perinatal mental health indicators are currently tracked routinely across countries in health information management systems (HIMS) or through standardized national health surveys as Demographic and Health Surveys (DHS) or Multiple Indicator Cluster Surveys MICS). Existing indicators, mainly from high-income countries, vary widely between countries and regions, making it challenging to compare data and assess the effectiveness of interventions [14, 15, 16].

Moreover, many indicators focus predominantly on the diagnosis and treatment of perinatal mental health conditions (mainly postpartum depression) rather than on the services provided and the access of women to those services, ensuring preventive measures and appropriate referral. There is also a need for more comprehensive and integrated approaches to perinatal mental health monitoring that address the complexity of healthcare systems and policies across countries ensuring integration across levels of care and within maternal, newborn and child health services [17]. Standardized indicators are essential for assessing the burden of perinatal mental health conditions globally and should show progress to address perinatal mental health issues, as well as guide policy and resource allocation.

### Overview of the Global PMH Theory of Change and scoping review on PMH indicators

A Global PMH Theory of Change (ToC) was published in 2022 [18] providing a strategic roadmap for developing standardized indicators for perinatal mental health monitoring. It outlines the underlying assumptions and pathways through which the proposed indicators are expected to bring about positive changes in the monitoring and management of perinatal mental health conditions. By identifying key inputs, activities, outputs, outcomes, and impacts, the ToC serves as a guide for understanding how the proposed indicators can contribute to improved maternal and newborn health outcomes. The ToC is based on a social ecological model where its domains are in order: individual, interpersonal relationship, community, service delivery ecosystem and policy landscape.

Further a scoping review conducted in 2023 aimed to identify existing perinatal mental health indicators used in monitoring systems worldwide [17]. The search in the scoping review was conducted in accordance with the Preferred Reporting Items for Systematic Reviews and Meta-Analyses Extension for Scoping Reviews. After consultation with WHO it was decided to exclude indicators related to risk factors for PMH or did not measure PMH directly. Additional indicators which measured mental health but were not specific to the perinatal period were excluded, as these are well described in WHO Mental Health Atlas [19]. Results were compiled and mapped to the relevant level in the Global PMH ToC. The review considered the indicators which: (1) aligned with the Global PMH ToC and landscape analysis definition of perinatal mental health (during pregnancy and up to one year after childbirth); (2) had a definition, including how it could be measured; and (3) was published from 1 January 2000 to 6 September 2023.

Nine indicators were identified which addressed individual (six indicators) and service-level (three indicators) characteristics (**Table 1**). In order to be reflective of global differences in the feasibility and availability of data to determine if indicators are met, stakeholder engagement in determining a final set of indicators is needed.

**Table 1.**
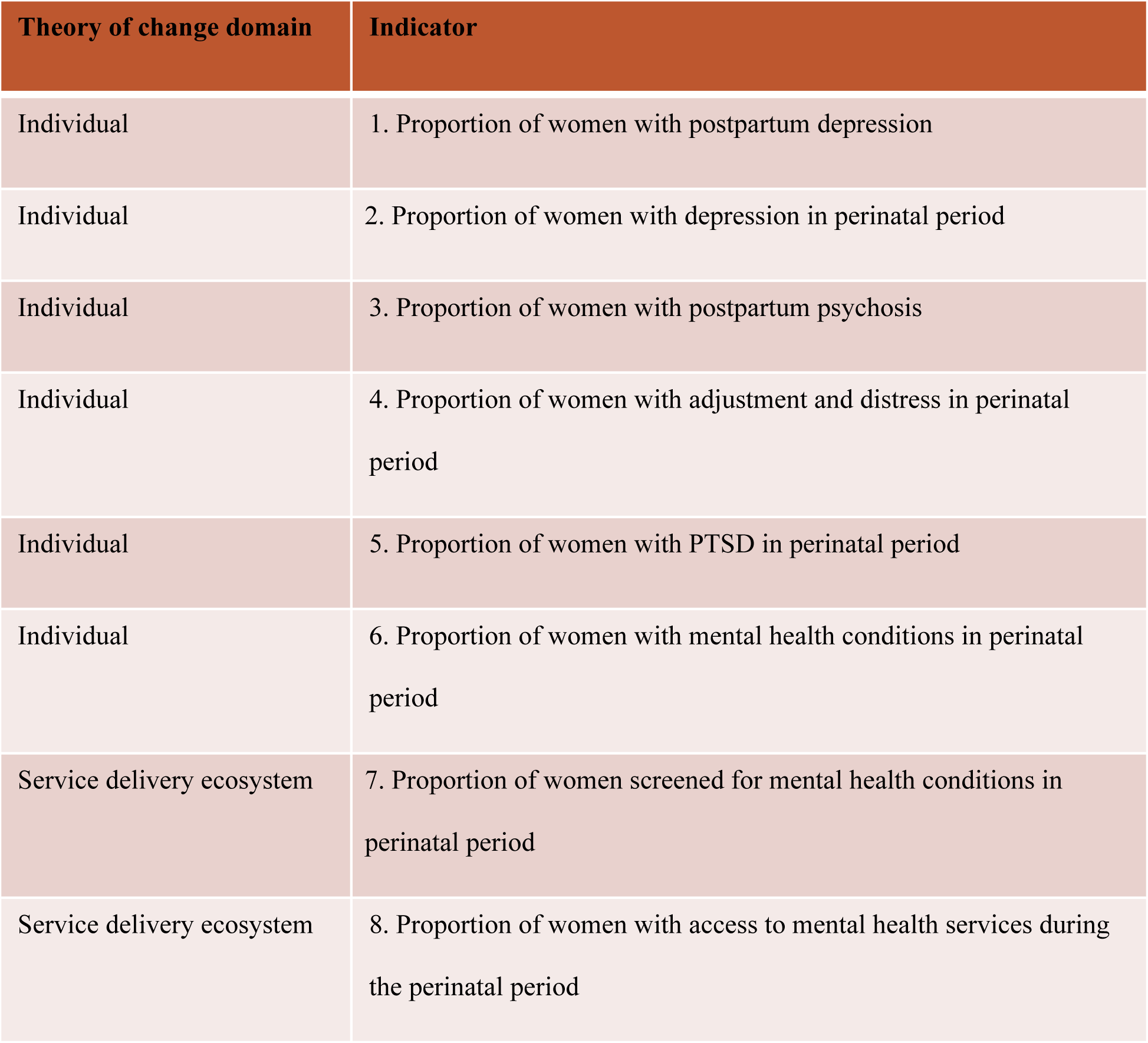
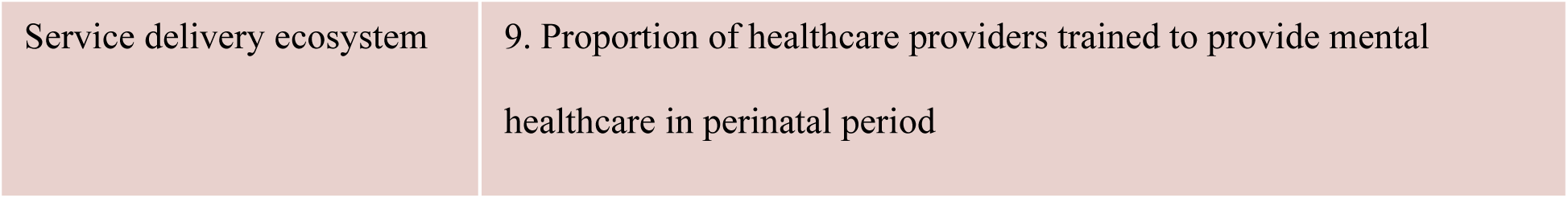
List of PMH indicators identified in the scoping review.

The purpose of this paper is to present the process for developing a set of primary and secondary indicators for countries in monitor PMH, discuss challenges in implementation of screening methods and training providers, and provide policy recommendations for integrating perinatal mental health indicators into existing monitoring frameworks.

## Methods

The development of a set of indicators for perinatal mental health involved a multi-stage process to ensure robustness and inclusivity of all World Health Organization (WHO) regions. This was a four-phase process (**Table 2)** guided by a WHO working group of experts and included online surveys based on the identified indicators of the scoping review and in consultation with the WHO Mother and Newborn Information for Tracking Outcomes and Results (MoNITOR) advisory group. This process brought to the finalization of global indicators on PMH with primary and secondary indicators for further reflection and adjustment.

**Table 2.** Phases to develop global PMH indicators.

| Phase 1 | Phase 2 | Phase 3: | Phase 4: |
| --- | --- | --- | --- |
| Establishment of WHO experts group on PMH monitoring and discussion on identified indicators of the scoping review | First survey with WHO experts group on PMH monitoring | Second survey with WHO experts group on PMH monitoring and additional suggested stakeholders from global, regional and country level | Consultation with WHO experts group on PMH monitoring and the WHO MoNITOR advisory group and development of global PMH indicators |

In phase one, a WHO working group of experts, comprised of 21 experts in perinatal mental health and monitoring and evaluation, from diverse regional backgrounds, was convened to provide guidance on indicator selection. The role of the WHO working group included providing feedback on the scoping review, on the list of proposed indicators and relevance in relation to the ToC; feedback and review on the findings from surveys, and of the proposed indicators, including feasibility of implementation.

In phases two and three, two rounds of surveys were conducted through an anonymized online form, among an international group of stakeholders, including healthcare professionals, researchers, policymakers, and representatives from civil society organizations. In phase two with the first survey, respondents were asked to select the top three PMH indicators for global monitoring from the list of nine indicators identified through the previous scoping review [17] and to propose any additional indicators used at the country-level which may be useful for global monitoring. During phase three the second survey included a wider group of stakeholders invited to order the list of indicators, selected from the previous survey, from the most to the least important indicator; and to propose additional ones if relevant. After each round of survey, the WHO working group of experts had online consultations to provide feedback on the findings. The specific questions of the two surveys are reported in **S1 First and second surveys**.

The first survey was completed by 21 members of the WHO working group, resulting in a total of 17 answers out of 21 participants. The second survey was completed by 35 stakeholders part of a broader group of experts involved more generally in maternal, newborn, child and adolescent health program implementation, mental health programme and monitoring and evaluation from global, regional and country levels. These stakeholders were identified through WHO regional offices and by the members of the WHO expert committee. The second survey requested participants to In the second survey the first question collected a total of 17 answers, while the second questions collected 8 answers.

Phase four included consultation with the WHO experts group on PMH monitoring and the WHO MoNITOR advisory group and development of global PMH indicators. Firstly, the feedback obtained from these surveys was analysed and a list of proposed indicators was developed to ensure their relevance and applicability across different settings. Later on, the WHO Mother and Newborn Information for Tracking Outcomes and Results (MoNITOR) advisory group and the WHO experts group on PMH monitoring, shared overall guidance and inputs on the results collected from the two surveys, which further informed the development process.

Inclusion and exclusion criteria were established to ensure the selection of indicators that met predefined quality standards and objectives. Indicators were included if they demonstrated validity, reliability, feasibility and potential impact for advocacy purposes. They also needed to be relevant to the assessment of perinatal mental health conditions and be able to measure progress towards improving maternal and newborn health outcomes for global monitoring purposes. Indicators were excluded if they lacked empirical evidence, were risk-related to develop PMH conditions or were overly complex or resource-intensive or were specific to a particular context and not generalizable to a broader population.

This work is based on the review of the literature and expert opinion, including WHO’s technical advisory groups on monitoring and measurement, and it is not original research. Therefore, we did not seek approval from the WHO ERC for ethical clearance.

We obtained written consent from members of the WHO expert group, following WHO protocols for expert group participation, which include declarations of interest and confidentiality agreements. The consent forms are securely stored on WHO servers, protected by a password. The WHO expert group has been involved in this activity on the prioritization of the PMH indicators from the 8th of December 2023 until the 25th of April 2024.

## Results

A summary of all steps in the selection of the indicators and results on the identification and prioritization of indicators can be found in **S2 Table.**

### Results from phase two (first survey)

A total of 17 out of 21 WHO Working Group members selected the top three PMH indicators for global monitoring choosing from the nine indicators identified through the previous scoping review [17] (**Table 3**).

**Table 3.** The PMH indicators for global monitoring in order of relevance by stakeholders from the first survey.

| **Theory of change domain** | **Indicator** | **Score** |
| --- | --- | --- |
| Service delivery ecosystem | 7. Proportion of women screened for mental health conditions in perinatal period | 12 (70%) |
| Service delivery ecosystem | 8. Proportion of women with access to mental health services during the perinatal period | 12 (70%) |
| Service delivery ecosystem | 9. Proportion of healthcare providers trained to provide mental healthcare in perinatal period | 11 (64%) |

Additionally, three more indicators have been suggested to be important for inclusion (although not classified as top three) which are at individual level: 2. “Proportion of women with depression in perinatal period”; 6. “Proportion of women with mental health conditions in perinatal period” and 1. “Proportion of women with postpartum depression”.

Nine respondents proposed additional indicators used at the country-level potentially useful for global monitoring:

- Postnatal care (PNC) disaggregated for women and baby
- Early Childhood Development Index (ECDI)
- System readiness indicators: 1. Presence of content on screening and counseling for maternal mental health in relevant pre-service curricula (MCH nurses; Community Health Workers etc.); 2. Antenatal care (ANC) and postnatal care (PNC) consultation /home visit guidelines including PMH screening, counseling & follow up
- Availability of PMH policy or included in national mental health policy
- PMH included in WHO country annual work plan
- PMH included in national health policy and other documents
- Proportion of health budget (mental health budget) allocated to maternal mental health (I have not used, but would be a useful indicator of high-level commitment at governance level)

### Results from phase three (second survey)

The survey requested 35 participants to put in order from the most important to the least the priority PMH indicators for global monitoring identified in the previous survey by the WHO working group of experts and to propose any additional indicator.

17 stakeholders completed the survey and the most voted indicators have been listed in order of priority:

1. Proportion of women with access to mental health services during the perinatal period;
2. Proportion of women screened for mental health conditions in perinatal period
3. Proportion of healthcare providers trained to provide mental healthcare in perinatal period;
4. Proportion of women with mental health conditions in perinatal period;
5. Proportion of women with depression in perinatal period
6. Proportion of women with postpartum depression

Additionally, eight respondents suggested the inclusion of additional indicators such as:

- Proportion of women with perinatal mental condition linked to psychosocial support OR mental health services
- Proportion of health budget allocated to perinatal mental health
- The proportion of women attending a first referral appointment for treatment of a mental health condition
- Women with anxiety during perinatal period and additional risk-related indicators for PMH

### Results from phase four

#### Results consultations with WHO expert committee

Following analysis of both surveys results, the key recommendations by the WHO expert committee were on the identification and prioritisation of indicators and on feasibility and implementation. These two aspects will be described separately.

##### Identification and prioritisation of indicators

There was consensus from the WHO expert group to include the list of indicators below based on prioritisation from stakeholders and likely feasibility:

- To include indicators number 1. “Proportion of women with postpartum depression” and 2. “Proportion of women with depression in perinatal period”, which are important although may be difficult to measure initially.
- To include indicator 6. “Proportion of women with mental health conditions in perinatal period”, and to better describe the common mental health conditions in the metadata.
- The committee also discussed the inclusion of indicator 7. “Proportion of women screened for mental health conditions in perinatal period” versus the possible inclusion of an indicator which measured the number of consultations on PMH through ANC and PNC services. It was decided to include indicator 7 since will already provide more specific information with the use of validated screening tools. Further, the WHO recommendations on maternal and newborn care for a positive postnatal experience currently include PMH services for PNC [20] but not for ANC [21], therefore different countries could have a different approach in providing PMH services.
- To include indicator 9. “Proportion of healthcare providers trained to provide mental healthcare in perinatal period”.

Then a list the indicators that were excluded (and rationale):

- To exclude indicator 8. “Proportion of women with access to mental health services during the perinatal period”. This indicator was excluded because of the unclear definition of “access”, and reliable measurement at a national level was considered less feasible.
- To exclude an indicator which measures anxiety during pregnancy although this can contribute to postpartum depression. The expert group acknowledged anxiety as an important issue in PMH and a precursor to and as a risk factor for depression. Since we were specifically excluding indicators of risk factors, this indicator has been excluded.

Additional new indicators that were added:

- The WHO expert committee recognized the importance of having two indicators for advocacy purposes on budget allocation and on existing national policies for PMH. If countries are unable to identify the budget specifically for PMH, the budget allocated to mental health may be reported instead
- To group some indicators as health system indicators: budget, trained staff, etc… Suggested grouping indicators by primary indicators to monitor from the start and additional secondary indicators to monitor within a later timeframe.

##### Feasibility and implementation

- To include the data source for each indicator to assist with prioritization. It was agreed to focus on a few indicators, define their purpose, and use them to advocate for services. It was recognised that PMH services may not be well defined, or even existent in many countries.
- To include recommendations consistently on screening tools to use for data collection, and where to locate resources and services at national level. Although screening is not universal, each country should have adopted a specific tool at national level.

#### Results from consultation with WHO MoNITOR advisory group

The consultation with WHO MoNITOR advisory group was focused on the feasibility and implementation of the global PMH indicators. They identified the need to refine the indicators related to perinatal mental health services, as well as the training and competencies required by healthcare providers in order to offer mental healthcare services during the perinatal period. It was also deemed essential to identify the specific mental health conditions that need to be measured and provide clear definitions for these conditions. Additionally, there should be a specification of the screening tools approved by WHO, along with the appropriate timeframe for their use. Clarification is needed on how and where to measure these indicators, along with considerations for ethical practices in the measurement process. The inclusion of referral processes and psychosocial support within the mental health services needs to be detailed. It is important to harmonize WHO guidelines by including perinatal mental health recommendations in guidelines on childbirth and antenatal care.

Given the variety of information to be collected and possible weaknesses in routine data collection systems, indicators could be collected by different sources. Some of the indicators could be integrated into HIMS or collected through national reporting (e.g. for indicators concerning existence of a national PMH policy, government expenditure, health providers trained), while others may be more reliably collected through national surveys like DHS or MICS or mental heath surveys (e.g. indicators on population prevalence of perinatal depression).

There should be a clear outline of the key elements required in a national plan for perinatal mental health, such as the tools to use, the planning period, training, and the types of services available. The risk of underreporting must be acknowledged. The indicator on the proportion of women with postpartum depression should be retained, as postpartum conditions are among the most frequently diagnosed. Finally, there should be specific screening for perinatal mental health conditions for women staying in the hospital for their small and sick newborns in NICUs.

#### Indicators for perinatal mental health monitoring

Based on the consultations a list of primary indicators and a list of secondary indicators was created based on capacity or quality data available (**Table 4** and **Table 5**). Metadata providing more detailed descriptions of each indicator is available in **S3 File**.

**Table 4.** Primary indicators for global monitoring.

| Monitoring domain | Indicator | Numerator | Denominator | Current preferred data source | Other data sources |
| --- | --- | --- | --- | --- | --- |
| Output | Proportion of countries with national policy /plan for perinatal mental health in SRMNAH or in the mental health policy/strategy | Number of countries with national policy for perinatal mental health in SRMNAH policy/strategy or in the mental health policy/strategy | Total number of countries | Policy review or WHO Policy survey |  |
| Outcome | Proportion of women screened for mental health conditions in perinatal period <sup>1</sup> | Number of women who were screened for mental health conditions in in perinatal period <sup>1</sup> | Total number of women who gave birth within a given time period | Household surveys | RHIS |
|  | Proportion of women with depression in perinatal period<sup>1</sup> | Number of women identified with depression<sup>2</sup> in the perinatal period<sup>1</sup> | Total number of women who gave birth within a given time period | Household survey | RHIS |

**Table 5.** Secondary indicators for global monitoring.

| Monitoring domain | Indicator | Numerator | Denominator | Current preferred data source | Other data sources |
| --- | --- | --- | --- | --- | --- |
| Output | Proportion of government allocated expenditure to perinatal mental health | Total government allocated expenditure to perinatal mental health | Total government allocated expenditure on health<sup>3</sup> |  |  |
|  | Proportion of in-service health care providers trained to provide mental healthcare services<sup>4</sup> in perinatal period | Number of in-service health care providers trained to provide mental healthcare services in perinatal period | Total number of in-service health care providers providing services in perinatal period | Health facility assessment | NHAs |
| Outcome | Proportion of women with perinatal mental health condition linked to psychosocial support OR mental health services | Number of women with perinatal mental health condition linked to psychosocial support OR mental health services | Total number of women with perinatal mental health condition | Household survey | RHIS |

1 Perinatal period: during pregnancy and up to 1 year after pregnancy
2 Depression following consultation and use of available screening tool
3 Relatively crude indicator
4 Mental Healthcare services include a variety of interventions, and it is not limited to screening of mental health conditions.
Please refer to the WHO Guide for integration of perinatal mental health in maternal and child health services. Geneva: World
Health Organization; 2022.

The primary indicators proposed for perinatal mental health monitoring represent measures aimed at capturing key aspects of maternal psychological wellbeing during pregnancy and up to one year postpartum. These indicators have been carefully selected based on their significance in assessing the population prevalence, policy and health system plans related to perinatal mental health conditions. They serve as foundational metrics for monitoring PMH conditions and policies aimed at improving maternal and newborn health outcomes. Primary indicators include measures in individual and healthcare system domains, such as the prevalence of antenatal and postnatal depression, rates of screening coverage for perinatal mental health conditions in the perinatal period, and the proportion of countries with national policy or plan for perinatal mental health.

In addition to the primary indicators, secondary indicators are proposed to guide future efforts and advancements in perinatal mental health monitoring within a later timeframe. These secondary indicators represent desired outcomes and areas for improvement that may require additional research, resources, and policy initiatives to achieve. They encompass measures aimed at enhancing the comprehensiveness, accuracy, and equity of perinatal mental health monitoring, as well as addressing emerging challenges and priorities in the field. Secondary indicators include individual, health service delivery and healthcare system domains, assessing the proportion of women receiving timely and appropriate treatment for perinatal mental health conditions, the proportion of in-service health care providers trained to provide mental healthcare services, and the budget expenditure allocated to improve this population issue.

By setting ambitious yet achievable goals for the future, secondary indicators pave the way for continued progress and innovation in perinatal mental health monitoring and ultimately contribute to better outcomes for women and newborns worldwide.

## Discussion

The proposed global PMH indicators has significant implications for global monitoring efforts at global and national levels. By establishing a standardized set of metrics, countries can systematically track and compare the prevalence and outcomes of perinatal mental health conditions across regions and over time. This allows for more accurate assessment of the burden of perinatal mental health disorders and identification of populations at greatest risk. Additionally, standardized indicators facilitate the evaluation of interventions and policies aimed at improving maternal and newborn health outcomes, enabling countries to identify effective strategies and allocate resources more efficiently. Moreover, by raising awareness and prioritizing perinatal mental health on the global agenda, the proposed indicators can catalyze action and advocacy to address this critical public health issue. The WHO expert committee’s advised to choose indicators to reflect key dimensions of perinatal mental health including prevalence, screening, policy and health system plans related to perinatal mental health. Priority was given to indicators that were evidence-based, feasible to measure, and relevant to global monitoring efforts.

Consideration was also given to indicators that could be easily integrated into existing health information systems and data collection tools as the ones used by hospital and households’ surveys to facilitate implementation and sustainability.

### Challenges in implementing standardized monitoring systems

The need to strengthen coordination and technical leadership to harmonize recommendations for improved measurement and monitoring of data related to maternal and newborn heath has been highlighted in previous publications [17, 18] and by the WHO MoNITOR Advisory group [22]. Despite the potential benefits, implementing standardized monitoring systems for maternal and newborn health and in this case for perinatal mental health specifically, poses several challenges. These include resource constraints, particularly in low- and middle-income countries, where healthcare infrastructure, funding, coverage and poor accessibility of PMH services may limit data collection. The integration of PMH into maternal and newborn health services including validated screening and diagnostic tools would be essential in the implementation of a monitoring system [23].

There are also technical challenges related to data collection, reporting mechanisms, the quality of data, and interoperability of health information systems which hamper disparities in monitoring efforts between high-income and low-income countries [24]. The national routine health information system is essential for decision-making, providing regular data on delivery and utilization of services and can be used for program planning and track progress towards national and subnational targets [25]. In an effort to include new indicators as the ones proposed in this paper, it is important to consider the use of household and health facility survey. Household surveys are the primary source of coverage data for LMICs [26], however not all interventions suggested by this framework can feasibly be tracked using this method.

Linking household and facility surveys would improve the coverage measurement, linking care-seeking data from household surveys with service provision data from health facility assessments provides a unique opportunity to produce measures of population coverage that account for service quality [27].

These indicators offer the opportunity to be integrated into the next version of DHS or MICS. Additionally, cultural and contextual factors may influence the acceptability and uptake of standardized monitoring protocols, requiring tailored approaches to implementation. Addressing these gaps is essential for improving the accuracy, comprehensiveness, and equity of perinatal mental health monitoring worldwide and requires sustained commitment from governments, healthcare providers, and international partners, as well as investment in capacity building and health system strengthening. Starting with primary indicators to then expand to secondary indicators would enable each national system to build capacity and advocate for an improved monitoring system in this area.

### Opportunities for collaboration and capacity building for healthcare workers

Collaboration and capacity building are essential for the successful implementation of standardized monitoring systems for perinatal mental health. Multisectoral collaboration involving government agencies, healthcare providers, academia, civil society organizations, and international partners can leverage expertise and resources to support monitoring efforts [28]. Capacity building initiatives, including task sharing and group problem solving therapy can empower health workers and users with the knowledge and skills needed to respond, screen, diagnose, treat or refer to another specialist, perinatal mental health conditions effectively [29, 30]. This includes training on culturally sensitive approaches to care, evidence-based interventions, and integration of mental health services into routine antenatal and postnatal care [23]. By fostering collaboration and investing in capacity building, countries can strengthen their healthcare systems and improve outcomes for women and newborns.

### Policy recommendations for integrating perinatal mental health indicators into existing monitoring frameworks

Integrating perinatal mental health indicators into existing monitoring frameworks requires policy support and coordination at the national and international levels. Policymakers should integrate primary and secondary PMH indicators into existing health information systems and monitoring frameworks for maternal and newborn health, ensuring data on maternal psychological wellbeing are routinely collected and reported. Additionally, policies should prioritize funding for mental health services and workforce development, ensuring that healthcare providers have the resources and support needed to address perinatal mental health effectively [31]. Finally, policies should promote collaboration across sectors and encourage the adoption of evidence-based practices to improve perinatal mental health outcomes. By incorporating perinatal mental health indicators into existing monitoring frameworks. Countries can enhance their capacity to address this critical public health issue and achieve better outcomes for women and newborns.

The proposed global indicators for perinatal mental health encompass a range of measures aimed at capturing key aspects of maternal psychological wellbeing during pregnancy and the postpartum period. By tracking these indicators, we can better understand the burden of perinatal mental health conditions, identify gaps in care, and assess the effectiveness of interventions. Ultimately, these indicators have the potential to improve maternal and newborn health outcomes by informing policy and practice and guiding resource allocation to areas of greatest need. Additionally, pilot testing of these global PMH indicators to finalize a PMH monitoring framework in different settings is needed to help identify any practical challenges or limitations, conduct data validation, and allow for further refinement and optimization before full-scale implementation.

## Conclusion

Given the significant impact of perinatal mental health on maternal and newborn health outcomes, there is an urgent need for global stakeholders to prioritize the provision and monitoring of perinatal mental health care. This includes governments, international organizations, healthcare providers, researchers, and civil society organizations. Global stakeholders are called upon to pilot the global PMH indicators into existing monitoring frameworks and programs, allocating resources for mental health services, and promoting collaboration and capacity building among healthcare workers. By working together to address perinatal mental health, we can ensure that all women receive the support and care they need to thrive during pregnancy and the postnatal period, ultimately leading to better outcomes for both women and newborns.

## Data Availability

All relevant data are within the manuscript and its Supporting Information files.

## Acknowledgements

We acknowledge the kind contribution of each member of the WHO expert group on PMH monitoring and the WHO MoNITOR advisory group.

## Notes

### Competing Interest Statement

The authors have declared no competing interest.

### Funding Statement

Yes

### Author Declarations

This work is based on the review of the literature and expert opinion, including WHO’s technical advisory groups on monitoring and measurement, and it is not original research and no original data were collected. Therefore, we did not seek approval from the WHO ERC for ethical clearance.

## References

1. The Lancet. Perinatal depression: a neglected aspect of maternal health. Lancet. 2023;402(10403):667

2. World Organization WH. WHO guide for integration of perinatal mental health in maternal and child health services. Geneva; 2022.

3. WHO. ICD 11 Reference Guide. Geneva; 2019

4. Howard LM, Molyneaux E, Dennis C-L, Rochat T, Stein A, Milgrom J. Non-psychotic mental disorders in the perinatal period. The Lancet. 2014;384(9956):1775-88.

5. Jones I, Chandra PS, Dazzan P, Howard LM. Bipolar disorder, affective psychosis, and schizophrenia in pregnancy and the post-partum period. The Lancet. 2014;384(9956):1789-99

6. Manolova G, Waqas A, Chowdhary N, Salisbury TT, Dua T. Integrating perinatal mental healthcare into maternal and perinatal services in low and middle income countries. BMJ. 2023;381:e073343.

7. World Health Organization. Maternal mental health n.d. [Available from: https://www.who.int/teams/mental-health-and-substance-use/promotion-prevention/maternal-mental-health

8. Herba CM, Glover V, Ramchandani PG, Rondon MB. Maternal depression and mental health in early childhood: an examination of underlying mechanisms in low-income and middle-income countries. Lancet Psychiatry. 2016;3(10):983–92.

9. Roddy Mitchell A, Gordon H, Lindquist A, Walker SP, Homer CSE, Middleton A, et al. Prevalence of Perinatal Depression in Low- and Middle-Income Countries: A Systematic Review and Meta-analysis. JAMA Psychiatry. 2023

10. Wang Z, Liu J, Shuai H, Cai Z, Fu X, Liu Y, Xiao X, Zhang W, Krabbendam E, Liu S, Liu Z, Li Z, Yang BX. Correction: Mapping global prevalence of depression among postpartum women. Transl Psychiatry. 2021 Dec 20;11(1):640. doi: 10.1038/s41398-021-01692-1. Erratum for: Transl Psychiatry. 2021 Oct 20;11(1):543. doi: 10.1038/s41398-021-01663-6. PMID: 34930896; PMCID: PMC8688482.

11. Gelaye B, Rondon MB, Araya R, Williams MA. Epidemiology of maternal depression, risk factors, and child outcomes in low-income and middle-income countries. Lancet Psychiatry. 2016;3(10):973–82.

12. McNab S DS, Gomez P, Bhatti A, Khadka N, Kenyi E. The Silent Burden: Common Perinatal Mental Disorders in Low- and Middle-Income Countries. Washington, DC.: USAID MOMENTUM.; 2021.

13. Stein A, Pearson RM, Goodman SH, Rapa E, Rahman A, McCallum M, et al. Effects of perinatal mental disorders on the fetus and child. Lancet. 2014;384 North American Edition (9956):1800–19.

14. Molenaar JM, Boesveld IC, Kiefte-de Jong JC, Struijs JN. Monitoring the Dutch Solid Start Program: Developing an Indicator Set for Municipalities to Monitor their First Thousand Days-Approach. International journal of integrated care. 2022;22(4):8

15. Centers for Disease Control and Prevention Chronic Disease Indicators U.S. Department of Health and Human Services; 2023 [Available from: https://www.cdc.gov/cdi/.

16. Meneses Navarro S, Serván-Mori E, Heredia-Pi I, Pelcastre B, Nigenda G. Ethnic Disparities in Sexual and Reproductive Health in Mexico After 25 Years of Social Policies. Sexuality Research and Social Policy. 2022;19:1–16.

17. Layton E., Mitchell A.R., Kennedy E., Moran A.C., Palestra F., Chowdhary N., McNab S., Homer C.S.E., Maternal Mental Health Matters: A scoping review of indicators for perinatal mental health, Plos- to be confirmed

18. McNab SE, Fitzgerald L, Stalls S, Bhatti AM, Doggett E, Ricca J, et al. A Shared Vision for Improving Perinatal Mental Health in Low- and Middle-Income Countries: A Theory of Change and Prioritized Implementation Research Questions. USAID, MOMENTUM; 2022.

19. Mental health atlas 2020. Geneva: World Health Organization; 2021. Licence: CC BY-NC-SA 3.0 IGO.

20. WHO recommendations on maternal and newborn care for a positive postnatal experience. Geneva: World Health Organization; 2022. Licence: CC BY-NC-SA 3.0 IGO

21. WHO recommendations on antenatal care for a positive pregnancy experience. Geneva: World Health Organization; 2016.

22. Moller AB, Newby H, Hanson C, Morgan A, El Arifeen S, Chou D, Diaz T, Say L, Askew I, Moran AC. Measures matter: A scoping review of maternal and newborn indicators. PLoS One. 2018 Oct 9;13(10):e0204763. doi: 10.1371/journal.pone.0204763. PMID: 30300361; PMCID: PMC6177145.

23. Manolova G, Waqas A, Chowdhary N, Salisbury TT, Dua T. Integrating perinatal mental healthcare into maternal and perinatal services in low and middle income countries. BMJ. 2023 May 23;381:e073343. doi: 10.1136/bmj-2022-073343. PMID: 37220917; PMCID: PMC10203867.

24. Kabue MM, Palestra F, Katwan E, Moran AC. Availability of priority maternal and newborn health indicators: Cross-sectional analysis of pregnancy, childbirth and postnatal care registers from 21 countries. PLOS Glob Public Health. 2023 Jan 5;3(1):e0000739. doi: 10.1371/journal.pgph.0000739. PMID: 36962773; PMCID: PMC10021477.

25. Mallick L, Temsah G, Namaste S, Dontamsetti T, and Wang W. Using Health Management Information Systems Data to Contextualize Survey-based Estimates of Fertility, Mortality, and Wasting: DHS Occasional Paper; No. 12. ICF; Rockville, Maryland, USA: 2020. Available from: https://dhsprogram.com/pubs/pdf/OP12/OP12.pdf. [Google Scholar]

26. Munos MK, Stanton CK, Bryce J, Core Group for Improving Coverage Measurement for MNCH Improving coverage measurement for reproductive, maternal, neonatal and child health: gaps and opportunities. J Glob Health. 2017;7:010801. doi: 10.7189/jogh.07.010801

27. Kanyangarara M, Chou VB, Creanga AA, Walker N. Linking household and health facility surveys to assess obstetric service availability, readiness and coverage: evidence from 17 low- and middle-income countries. J Glob Health. 2018 Jun;8(1):010603. doi: 10.7189/jogh.08.010603. PMID: 29862026; PMCID: PMC5963736.

28. Ng’oma M, Bitew T, Kaiyo-Utete M, Hanlon C, Honikman S, Stewart RC. Perinatal mental health around the world: priorities for research and service development in Africa. BJPsych International. 2020;17(3):56–59. doi:10.1192/bji.2020.16

29. J E M Nakku, O Nalwadda, E Garman, S Honikman, C Hanlon, F Kigozi, C Lund. Group problem solving therapy for perinatal depression in primary health care settings in rural Uganda: an intervention cohort study (2021), BMC Pregnancy and Childbirth 21:584. DOI: 10.1186/s12884-021-04043-6

30. S Boisits, Z Abrahams, M Schneider, S Honikman, D Kaminer and C Lund. Developing a task-sharing psychological intervention to treat mild to moderate symptoms of perinatal depression and anxiety in South Africa: a mixed-method formative study (2021), Int J Ment Health Sys*t15*, 23. DOI: 10.1186/s13033-021-00443-5

31. Schiller CE, Cohen MJ, O’Hara MW. Perinatal mental health around the world: priorities for research and service development in the USA. BJPsych Int. 2020 Nov;17(4):87–91. doi: 10.1192/bji.2020.15. PMID: 33196694; PMCID: PMC7609989.

